# Brief remote psychological treatments for healthcare workers with emotional distress during the SARS-CoV-2 pandemic: a randomized clinical trial

**DOI:** 10.1101/2024.09.04.24313084

**Authors:** Giovanni Abrahão Salum, Marianna de Abreu Costa, Lucas Spanemberg, André Rafael Simioni, Natan Pereira Gosmann, Lívia Hartmann de Souza, Pim Cuijpers, Daniel Samuel Pine, André Russowsky Brunoni, Christian Haag Kristensen, Marcelo Pio de Almeida Fleck, Gisele Gus Manfro, Carolina Blaya Dreher

## Abstract

**BACKGROUND:** The SARS-CoV-2 pandemic has catalyzed a widespread mental health crisis, impacting millions of people. This study aimed to compare three brief remote psychological treatments for healthcare workers with emotional distress during the SARS-CoV-2 pandemic.

**METHODS:** Nationwide three-arm randomized clinical trial in Brazil. This is a transdiagnostic study that included professionals and students from health services with high levels of anxiety, depression, or irritability symptoms, as defined by Patient-Reported Outcomes Measurement Information System (PROMIS). The exclusion criterion was positive suicide risk. Participants were randomized single session psychoeducation group plus weekly personalized pre-recorded videos for four weeks (SSI-ET), brief cognitive behavioral telepsychotherapy group (B-CBT, four sessions), or brief interpersonal telepsychotherapy (B-IPT, four sessions). This study was registered in clinicaltrials.gov (NCT04635618). The primary outcome was the proportion of participants with a 50% reduction in T-scores in PROMIS rating scales of anxiety, depression, and/or irritability at one-month.

**FINDINGS:** Of the 3328 volunteers assessed for eligibility, 999 participants were enrolled, from May 19^th^ 2020 to December 31^st^ 2021, and allocated to SSI-ET (n=342), B-CBT (n=323), or B-IPT (n=334). All three groups showed significant symptom reductions in the one-month assessment that were maintained over the three and six-month follow-ups (Cohen’s d range = 0.94-1.36, p<0.001), with no significant difference between groups. The estimated proportion of responders were 46.5%, 43.7% and 44.6% for SSI-ET, B-CBT and B-IPT, respectively.

**Conclusions:** Our results refute the hypothesis that therapeutic interventions with higher number of sessions and with more specialized therapeutic components offer advantages in alleviating emotional distress, particularly among healthcare workers facing epidemic emergencies. These results have critical implications for planning interventions for crisis responses, especially in settings with limited resources.

**FUNDING:** Ministry of Health of Brazil, Coordenação de Aperfeiçoamento de Pessoal de Nível Superior, Conselho Nacional de Desenvolvimento Científico e Tecnológico, and Fundo de Incentivo à Pesquisa/Hospital de Clínicas de Porto Alegre.

## INTRODUCTION

The severe acute respiratory syndrome coronavirus 2 (SARS-CoV-2) pandemic triggered a new mental health crisis since its outbreak.^1^ Millions of people have faced mental health stressors including exposure to infection, bereavement, physical distancing, quarantine, and economic losses.^2^ Brazil has been severely hit by COVID-19, with more than 37 million cases and more than 702,000 deaths related to COVID-19.^3^ Among the different occupational groups, frontline healthcare workers had been especially at risk of adverse mental health consequences, due to the higher risk of infection, work overload, and psychological demands posed by their patients.^4^

Psychological treatments such as cognitive behavioral therapy (CBT) and interpersonal psychotherapy (IPT) have been shown to significantly reduce symptoms of anxiety, depression, and irritability in the adult population.^4^ Evidence of efficacy extends to guided forms of internet-delivered CBT,^5,6^ and a growing number of studies have demonstrated the effects of these psychotherapies in very brief online interventions.^7^ However, the length of psychotherapy for those exposed to traumatic events such as first responders is still in debate. On one hand, brief CBT was the only intervention that showed clinical improvement over an active treatment for those with trauma related symptoms according to meta-analyses and recent guidelines.^8–10^ On the other hand, the ‘psychological debriefing’,^9,10^ a one session intervention that was developed for those exposed to trauma, was no longer recommended for being, at best, ineffective.^10^ Although a recent meta-analysis claims that this recommendation relies on studies with high heterogeneity,^10^ the current state of art for first responders recommends minimal interventions for those exposed to trauma (psychological first aid) to brief interventions for those with acute stress response.^8^ These recommendations focus on trauma related symptoms, although anxiety was more frequently presented in frontline healthcare workers.^8^ Besides this, studies have also shown that the effects of these psychotherapies are not limited to diagnostic-specific approaches, but can also benefit dimensional approaches for people with multiple emotional problems.^8^

While the efficacy of these brief remote interventions is well established in clinical scenarios, little is known about their utility in crisis responses, such as the SARS-CoV-2 pandemic. In crisis situations, brief interventions such as ‘Psychological First Aid’, have been widely recognized as the first line choice for providing care,^11^ despite scarce evidence. Finally, the integration of these techniques with routine outcome assessment and risk stratification, basis for enhanced psychoeducation, is yet to be examined. Therefore, little is known about how to help people highly exposed to crisis situations, and whether the addition of more complex interventions, such as CBT or IPT, in brief formats can add to an enhanced single-session psychoeducation intervention that includes principles of Psychological First Aid, routine outcome assessment, and risk stratification.

This study aimed to compare three brief remote psychological treatments for health workers with emotional distress (anxiety, depression, or irritability) during the SARS-CoV-2 pandemic. We hypothesize that a brief four-week protocol of cognitive behavioral telepsychotherapy (B-CBT) and a brief four-week protocol of interpersonal telepsychotherapy (B-IPT) will increase the proportion of individuals experiencing symptom reduction as compared to a single session intervention enhanced telepsychoeducation (SSI-ET), and that there will be no superiority between B-IPT and B-CBT.

## METHODS

### Study design

The TelePSI trial is a nationwide, investigator-initiated, unicenter, randomized, unblinded, pragmatic controlled trial conducted in Brazil. This trial aimed to evaluate the effectiveness of three different remote interventions on transdiagnostic psychiatric symptoms: SSI-ET, B-CBT, and B-IPT. The trial was designed and supervised by a steering committee of members from academia and international experts in emotional disorders (DSP) and psychotherapy for emotional disorders (PC).

The trial protocol was approved by the National Research Ethics Commission (CONEP). This article adhered to the ethical principles outlined in the Declaration of Helsinki and to guidelines established in the approved protocol and is reported as recommended by CONSORT 2010 guideline (appendix p 2). This trial received funding from the Ministry of Health of Brazil through the decentralized execution term No. 16/2020.

### Participants

Participants were recruited nationwide via helplines, e-mails, social media, and traditional media (e.g., national and local newspapers and TV commercials). All assessments and study procedures were conducted online and all participants provided oral informed consent.

The inclusion criteria were the following: professionals or interns from the health sector and a T-score of 70 or above in any of the Patient-Reported Outcomes Measurement Information System (PROMIS) scales of anxiety, depression or anger.^12^ The exclusion criterion was suicidal ideation assessed using one question from the Patient Health Questionnaire (PHQ-9).^13^ Individuals who rated one, two, or three on this question were immediately referred to a psychiatrist session to ensure participants’ safety.^14^ All participants that underwent the psychiatric assessment were offered psychotherapy after being cleared for the suicide risk, but were excluded from this analysis. During all the follow-up, the therapist could referee the participants to a psychiatric evaluation whenever a risk was identified. A total of 16 participants were referred to psychiatric assessment during the trial (three in the B-CBT group, eight in the SSI-ET group, and six in the B-IPT group). In all 16 cases, risk was absent or mild, not requiring any additional measure or interventions. There was no restriction for inclusion regarding self-reported sex at birth (i.e., male or female). The demographic characteristics of the sample are depicted in table 1 and appendix (p 10).

**Table 1.** Sample description among groups.

|  | **SSI-ET (n=342)** | **B-CBT (n=323)** | **B-IPT (n=334)** |
| --- | --- | --- | --- |
| **Sex at birth (female)** | 307 (90%) | 290 (89.8%) | 293 (87.7%) |
| **PROMIS Anxiety** |  |  |  |
| None to slight | 0 (0%) | 0 (0%) | 0 (0%) |
| Mild | 0 (0%) | 1 (0.3%) | 1 (0.3%) |
| Moderate | 26 (7.6%) | 23 (7.1%) | 30 (9.0%) |
| Severe | 316 (92.4%) | 299 (92.6%) | 302 (90.4%) |
| **PROMIS Depression** |  |  |  |
| None to slight | 20 (5.8%) | 19 (5.9%) | 12 (3.6%) |
| Mild | 70 (20.5%) | 60 (18.6%) | 51 (15.3%) |
| Moderate | 206 (60.2%) | 208 (64.4%) | 213 (63.8%) |
| Severe | 46 (13.5%) | 36 (11.1%) | 58 (17.4%) |
| **PROMIS Irritability** |  |  |  |
| None to slight | 35 (10.2%) | 35 (10.8%) | 26 (7.8%) |
| Mild | 39 (11.4%) | 35 (10.8%) | 32 (9.6%) |
| Moderate | 152 (44.4%) | 153 (47.4%) | 159 (47.6%) |
| Severe | 116 (33.9%) | 100 (31%) | 117 (35%) |
| **Age (years) - Mean (SD)** | 35.3 (9.31) | 36.0 (8.91) | 35.2 (8.64) |
| **PROMIS Anxiety (T-score) - Mean (SD)** | 73.4 (3.06) | 73.5 (3.43) | 73.6 (3.86) |
| **PROMIS Depression (T-score) - Mean (SD)** | 63.7 (5.82) | 63.6 (5.56) | 64.8 (5.51) |
| **PROMIS Irritability (T-score) - Mean (SD)** | 66.5 (8.74) | 66.5 (9.11) | 67.6 (8.69) |
| **Satisfaction with Life (sum) - Mean (SD)** | 16.7 (6.01) | 17.6 (5.57) | 17.3 (5.96) |

PROMIS, Patient-Reported Outcomes Measurement Information System; SSI-ET, single session intervention - enhanced psychoeducation; B-CBT, brief cognitive behavioral telepsychotherapy; B-IPT, brief interpersonal telepsychotherapy; SD, standard deviation.

### Randomisation and masking

Participants were randomly allocated in a 1:1:1 ratio to SSI-ET, B-CBT, or B-IPT. Randomization occurred at the intervention and therapist levels immediately after the participants completed the online questionnaires and was centrally conducted by independent research assistants. An automated computer-generated random numbers table was used to perform the randomization. It automatically assigned participants to one of the three arms based on their trial ID numbers. Due to the nature of the interventions, both participants and therapists were aware of the assigned intervention.

### Procedures

The treatments were delivered by clinical psychologists or psychiatrists with previous training in each clinical approach. The interventions were performed by eight therapists in SSI-ET and B-IPT arms and by six therapists in the B-CBT arm. All therapists underwent an online training for the specific protocols and attended to weekly or biweekly supervisions during the trial. The length of the protocol training varied from three to eight hours and included one video of each session and an online test.

The first psychotherapy session started no later than one week after randomization. All psychotherapies were conducted online using Google Meet. At the end of the treatment (week four), we sent online self-report questionnaires to the participants via phone. Follow-up assessments were conducted three months and six months after treatment and the participants received detailed reports of their answers via email. In cases where respondents did not respond to the questionnaire links, the research team sent up to six reminders to encourage completion of the assessments.

The SSI-ET protocol was constructed considering the best available evidence for dealing with crisis situations and supporting the treatment of anxiety, depression, and irritability. Regarding crisis situations, our protocol was largely based on the principles of Psychological First Aid. For the treatment of anxiety, depression, and irritability, we relied on extensive evidence showing the importance of psychoeducation and routine outcome assessment (measurement-based care). This intervention was created specifically for this study. The training for this SSI involved a detailed manual with examples on how to conduct the intervention, online lectures explaining the intervention, simulated sessions, and a questionnaire assessing knowledge acquisition. All therapists underwent weekly supervision sessions to guarantee the fidelity of the interventions. This protocol consisted of a one-session intervention with a trained psychologist which sent two brief pre-recorded videos per week for four weeks based on the needs identified in the synchronous session. Therapists were instructed to carefully and empathetically listen to the participants, and create a welcoming, gentle, and non-judgmental environment in which participants felt comfortable and confident to express their thoughts, concerns, and emotions.^15^ The general principle was assisting participants in stressful situations, encouraging them to develop autonomy, self-efficacy, and problem-solving without harming, strengthening their support network, and developing the ability to feel safe, not alone, calm, and hopeful, as well as to ensure social, physical and emotional support. During the session, conducted by qualified psychologists or psychiatrists, the therapist would first review participants’ symptom levels using scores obtained from PROMIS Anxiety, Depression and Anger scales. Soon after, the participant’s responses regarding protective behaviors (e.g. meditating, reading books, eating healthily) and risky behaviors (e.g. self-medication, excessive alcohol consumption, problematic use of social media) were reviewed and the participant encouraged to adopt safe coping mechanisms. Both the PROMIS scale scores and the survey of risk/protective behaviors were shared in the form of visually appealing graphics during the online session.

To personalize the treatment, psychoeducation videos were carefully selected based on participants’ responses to PROMIS scales, risk/protective behaviors, and their expressed needs during the session. Over the following four weeks, the therapists sent a total of eight videos (two videos per week) to the participants. These videos were selected from a list of 16 that covered the following topics: contagion fear, typical vs. excessive anxiety, typical vs. excessive depression, anger vs. irritability, burnout, acute stress reactions, sleep hygiene, healthy eating habits, exercise, excessive consumption of alcohol and drugs, excessive exposure to news, excessive use of social media, taking care of children, taking care of the elderly, and social support. These videos were developed by the researchers and are freely available on TelePSI website (telepsi.hcpa.edu.br). Additionally, the therapists were available for four weeks to address any messages or queries sent by the participants through a phone chat. Given therapists were instructed to keep direct contact with participants to a minimum, additional contact via this platform was reported to be minimal. The appendix (pp 4-5) presents additional information regarding session format, therapist techniques, and content of the videos.

The B-CBT was a four-session intervention focusing on emotional symptoms. This strategy aimed to provide resources for health care professionals to cope with stress, anxiety and depressive symptoms. The first session was similar to the SSI-ET, including empathetic listening, feedback on clinical scales with psychoeducation, identification on protective and risky behavior, encouragement on self-efficacy and adopting protective coping mechanisms. Besides this, this session included CBT psychoeducation and setting the follow-up appointments. The three following sessions of this protocol follows a transdiagnostic approach based on the Unified Protocol for Transdiagnostic Treatment (UP), an intervention designed to address neuroticism, the core temperament associated with anxious and depressive disorders.^16^ Due to the pandemic context, we also included techniques to cope with insomnia, acute stress, emotional distress, and irritability. Each session was followed by two booster videos on CBT techniques sent through text messages. Each session lasted for one hour and was conducted by qualified psychologists or psychiatrists specialized in CBT, being followed by two booster videos sent through text messages. The videos selected during the therapy sessions included those available in the psychoeducation group (for the first week), as well as six specific videos tailored to CBT: diaphragmatic breathing, mindfulness, cognitive flexibility, cognitive model, problem-solving and behavioral activation. The appendix (pp 6-7) presents a comprehensive explanation of the intervention structure and content.

The B-IPT is a four-session adaptation of interpersonal therapy based on The Edinburgh Early Intervention Model: Psychological First Aid & Abbreviated Interpersonal Psychotherapy adapted for COVID-19^17^ and Interpersonal Counseling (IPC) for depression in primary care.^18^ IPT, known for its effectiveness in crisis contexts, focused on problem areas related to grief, interpersonal disputes, and role transitions. The first session was similar to the SSI-ET, including empathetic listening, feedback on clinical scales with psychoeducation, identification on protective and risky behavior, encouragement on self-efficacy and adopting protective coping mechanisms. Besides this, the first session included IPT psychoeducation, interpersonal inventory assessment and identification of problem areas that will be explored during the further three sessions of treatment (grief, interpersonal disputes, and role transition) and setting the follow-up appointments. The problem area elected to be the focus of treatment in sessions two and three, the initial symptoms of emotional distress are reviewed, relating them to the interpersonal context. The problem area identified in session one will be addressed in subsequent sessions, maintaining continuity between sessions and deepening strategies for dealing with the focused situation. The therapist may utilize the full range of TIP techniques, such as non-directive exploration, directive elicitation, clarification, communication analysis, decision analysis, role-playing, advice-giving, and mentalization. In session four, in addition to the usual symptom review, the termination of treatment is explicitly discussed. The therapist helps the participant recognize their competence through reviewing the interpersonal network, revisiting the course of treatment and the participant’s progress, and assessing early warning signs, discussing future alternatives.

The therapy sessions were conducted by psychologists and psychiatrists specialized in IPT through one-hour video calls. Two videos were sent each week. The videos selected for the therapy sessions included those available in the psychoeducation group (sent in the first week), as well as specific videos tailored to IPT that include how to communicate with others, how to ask for help, grief, encouraging the expression of feelings, interpersonal disputes, and role transitions. The appendix (pp 8-9) presents a comprehensive description of the intervention structure and content.

The baseline assessment was completed online by the participant before the randomization. The therapist contacted and scheduled the first appointment within one week after the baseline assessment. The following appointments were delivered during the next four weeks and could not exceed six weeks after the baseline assessment. The one-month assessment was performed 30 days after the first session as long as the participant completed the four-weeks protocol. The follow-up assessments were performed 3 months and 6 months after the first session.

### Outcomes

The primary outcome was the proportion of participants with a 50% reduction in T-scores on the Patient-Reported Outcomes Measurement Information System (PROMIS) scales of anxiety, depression, and/or irritability at one-month. The secondary outcomes were remission rates at months one, three, and six assessed through the proportion of patients with a T-score of 50 or below in all three emotional distress PROMIS subscales, and mean score changes between groups in anxiety, depression, and irritability. Other secondary outcomes were life satisfaction, measured with Satisfaction with Life Scale, which captures individuals’ appraisal of their overall well-being and contentment with life and reports its results with higher scores indicating higher satisfaction with life., and service satisfaction assessed through net-promoter score (NPS), calculated by asking participants a single question regarding the likelihood of recommending each therapy to someone on a scale from 0 to 10. Respondents were classified as promoters (9-10), passives (7-8), or detractors (0-6). The NPS was derived by subtracting the percentage of detractors from promoters. We also asked participants to rate their overall satisfaction using a single question ranging from 1 (’Extremely dissatisfied’) to 7 (’Extremely satisfied’). Measures of anxiety, depression, irritability (i.e. the primary outcomes), and life satisfaction were measured at months one, three, and six, and service satisfaction was evaluated at month one.

Participants’ safety was ensured by screening suicidal ideation through the Suicide item of the Patient Health Questionnaire (PHQ-9): ‘Over the last two weeks, how often have you been bothered by thoughts that you would be better off dead or of hurting yourself in some way?’. The response options are ‘Not at all’, ‘Several days’, ‘More than half days’, and ‘Nearly every day’.

Therapists’ fidelity to each protocol was assessed by a supervisor who reviewed a series of recorded sessions that were randomly selected from each therapist. The supervisor used a scale developed by our team to evaluate the quality and fidelity of each session. In addition, the supervisor met the therapists in group supervision weekly to ensure protocol fidelity. The assessment of the session quality involved 12 distinct criteria. Among these, seven skills are overarching and common to various interventions: (1) establishing a therapeutic alliance with the patient; (2) structuring and organizing the session; (3) compassion, empathy, genuineness, and warmth; (4) effective use of non-specific therapeutic skills such as clarification, reflection, and validation; (5) confidence and self-assurance; (6) reassurance; and (7) proficiency in instilling hope. For therapists practicing SSI-ET, assessment focused exclusively on general skills. An additional set of five skills pertains specifically to CBT: (1) overall adherence to a CBT theoretical orientation, (2) initiating sessions with receptiveness to the patient’s requests, (3) establishing a clear agenda for each session, (4) utilization of summaries and feedback throughout the session, and (5) use of the cognitive model. In contrast, the five additional skills for IPT were the following: (1) overall alignment with the IPT techniques, (2) discussion of the patient’s role as an active participant in the recovery process, (3) focus on a specific problem area throughout the sessions, (4) addressing symptoms of emotional distress within the interpersonal context, and (5) exploring strategies to enhance social support. Each item was rated on a scale of 1 (low) to 5 (high), with higher scores indicating greater session fidelity.

### Choice of primary outcome

The primary outcome was the proportion of participants with a 50% reduction in T-scores on PROMIS scales of anxiety, depression, and/or irritability at one-month. PROMIS is a self-rated assessment tool developed by the National Institutes of Health (NIH) to assess patients’ perspectives on their health status and overall quality of life. These scales, covering emotional states over the previous seven days, comprise 8 items for anxiety and depression each, and 5 items for anger. Responses are recorded on a 5-point Likert scale (1=’Never’, 2=’Rarely’, 3=’Sometimes’, 4=’Often’, 5=’Always’), with higher scores indicating increased emotional distress. All PROMIS scales have high internal consistency and reliability.^12^ We used PROMIS T-scores with an average of 50 and a standard deviation of 10. Therefore, a T-score of 70 (i.e., our inclusion criterion) represents 2 standard deviations above the sample mean. This measure was chosen as the primary outcome since this instrument is designed to be patient-friendly and uses fewer items than most scales designed to evaluate the same symptom domains, thereby decreasing respondent burden. Also, this instrument is freely available and has been translated into several languages.

### Statistical analysis

The study was designed to have 90% power to detect a 15% difference between groups in the primary outcome (alpha 0.017 [3 comparisons 0.05/3]). Estimating a 20% loss to follow-up, we needed a total of 999 participants. Because the enrollment rate was lower than expected in the first year, and the pandemic extended for a long time, the steering committee decided to prolong enrollment until the sample size was achieved.

Analysis was performed according to intention to treat criteria (ITT). Prior to the analysis of categorical outcomes, we performed a multiple imputation procedure to account for missing data. Multiple imputations were performed using as predictors the main outcomes for response and remission in each time point (endpoint, 3-months and 6-months) and the PROMIS classification for the baseline assessments. These imputations were conducted using 50 imputed datasets and ten iterations. All summary statistics and predictions were conducted using pooled estimates across all the 50 imputed datasets. Rates of response and remission at all time points were assessed using a combination of chi-square statistics for multiply imputed data. NPS and service satisfaction were also tested in the imputed dataset using general linear models.

Continuous outcomes were analyzed using linear mixed models with all available data from baseline, one, three and six-months follow-up. This analysis was performed in the non-imputed dataset and considered maximum likelihood estimation. Two-sided p-values of 0.05 or less were considered to indicate statistical significance in omnibus tests, and a p-value of 0.017 were considered significant for pairwise post-hoc comparisons.

In order to formally test for the absence of minimal clinically relevant differences between treatment arms, we have also included, *a posteriori,* an equivalence test (two one-sided tests equivalence testing, TOST) between interventions considering −15% as the smallest effect size of interest, as defined *a priori* for the statistical power analysis.^19^

All analyses were performed using R. Multiple imputation, combined chi-square statistics, linear mixed models, equivalence tests, and parameters were conducted or estimated using *Mice*, *Miceadds*, *lme4*, *TOST*, and *Effects* packages.^20–22^ Standardized mean differences (SMD) or Cohen’s *d* were used as a measure of standard effect size for significant associations, with values of 0.2, 0.5, and 0.8 interpreted as small, medium, and large, respectively.

Due to the crisis context of the SARS-CoV-2 pandemic, this trial was prospectively registered at ClinicalTrials.gov, NCT04635618 in November 11^th^ 2020, during the recruitment process; nevertheless, less than 6% of participants had already been included by the time of registration, no preliminary statistical analyses were performed, and almost all methodological procedures had already been defined and were followed throughout the entire trial. The sole deviation from the protocol is that, rather than excluding participants based on a clinical assessment of suicide, we excluded them based on affirmative responses to the PHQ-9 suicide item. This decision stemmed from the concern that randomizing participants after the psychiatric assessment might introduce bias in a trial designed to compare distinct interventions against a single session.

### Role of the funding source

The funders of the study had no role in study design, data collection, data analysis, data interpretation, writing of the report, and decision to submit the article for publication.

## RESULTS

Of the 3328 individuals who sought to participate in this project from May 19^th^ 2020 to December 31^st^ 2021, 3086 consented to participate in the study, and 490 were excluded and subsequently referred to a psychiatrist due to concerns about suicide risk. We enrolled 999 participants who met the inclusion criteria, being subsequently allocated to SSI-ET (n=342), B-CBT (n=323), or B-IPT (n=334) (figure 1). The majority of the sample consisted of women (89.2%), and the mean age of the entire sample was 35.5 (SD=8.9). Each group is further described in table 1 and appendix (p 10).

**Figure 1.**
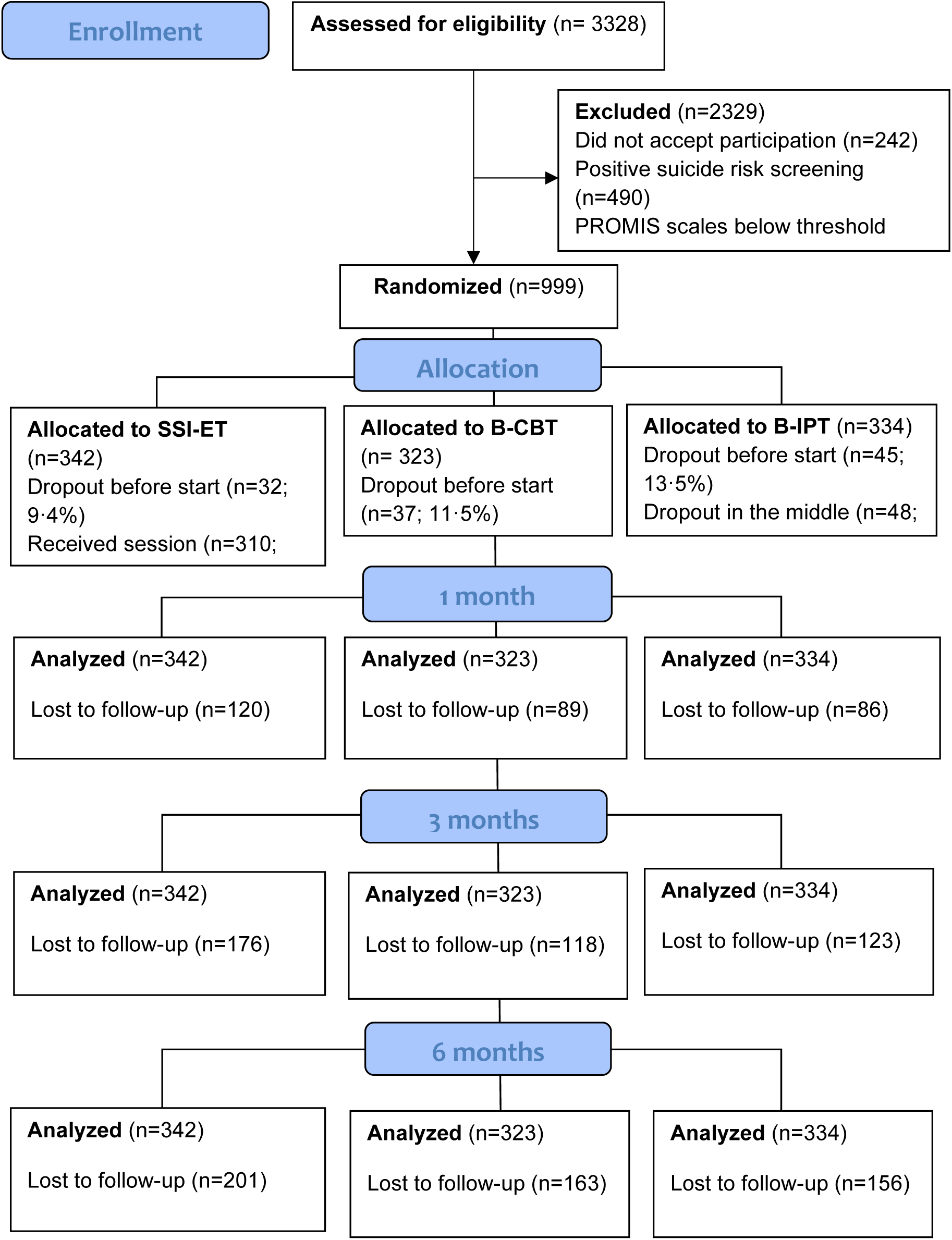
CONSORT Flow Diagram

### Primary outcome (one-month)

We did not find statistically significant between-group differences in the primary outcome with estimated proportion of responders being 46.5%, 43.7%, and 44.6% for SSI-ET, B-CBT, and B-IPT, respectively (combined χ^2^, F[2,1304.8]=0.315, p=0.729). Pairwise contrasts revealed no significant differences between SSI-ET vs. B-CBT (OR=1.12,95% CI 0.77-1.61, p=0.540), SSI-ET vs. B-IPT (OR=1.03, 95% CI 0.72-1.48, p=0.837), and B-CBT vs. B-IPT (OR=0.92, 95% CI 0.64-1.33, p=0.682).

### Response rates (three-month and six-month follow-up)

Estimated proportion of responders for three-months follow-up were 41.5%, 44.6%, and 41.1% for SSI-ET, B-CBT, and B-IPT, respectively (combined χ^2^, F[2,433.4]=0.412, p=0.662). Pairwise contrasts revealed no significant differences between SSI-ET vs. B-CBT (OR=0.879, 95% CI 0.58-1.33, p=0.541), SSI-ET vs. B-IPT (OR=0.86, 95% CI 0.58-1.28, p=0.462), and B-CBT vs. B-IPT (OR=0.98, 95% CI 0.68-1.42, p=0.926). There were also no significant differences for the six-months assessment, with estimated proportion of responders being 40.1%, 42.9%, and 33.4% for SSI-ET, B-CBT, and B-IPT, respectively (combined χ^2^, F[2,245.3]=2.079, p=0.127). Contrasts revealed no significant differences between SSI-ET vs. B-CBT (OR=0.91, 95% CI 0.55-1.49, p=0.703), SSI-ET vs. B-IPT (OR=0.66, 95% CI 0.43-1.03, p=0.073), and B-CBT vs. B-IPT (OR=0.73, 95% CI 0.47-1.14, p=0.171).

### Remission rates

Remission rates were also not significantly different between groups: (a) 13.3%, 9.5% and 11.8% showing remission in the SSI-ET, B-CBT and B-IPT, respectively, in the one-month assessment (combined χ^2^, F[2,1304.9]=0.315, p=0.730); (b) 11.9%, 14.8% and 12.0% in the three-month assessment (combined χ^2^, F[2,289.5]=0.534, p=0.587), and (c) 13.2%, 16.5% and 11.1% in the six-month assessment (combined χ^2^, F[2,207.59]=1.231, p=0.294). None of the contrasts revealed any significant difference between groups (all p-values>0.05).

### Relevant time effects

Linear mixed models, utilizing all available data, showed prominent time effects for all outcomes, meaning significant differences from baseline to one, three, and six-month follow-up assessments (figure 2). Mean changes were strongly significant for each treatment arm. Standardized symptom reduction for anxiety symptoms from baseline to one-month was SMD=1.15 (95% CI 1.03 - 1.27, p<0.001) for SSI-ET, SMD=1.35 (95% CI 1.24 - 1.47, p<0.001) for B-CBT, and SMD=1.36 (95% CI 1.23 - 1.49, p<0.001) for B-IPT. Standardized symptom reduction for depressive symptoms from baseline to one-month was SMD=0.94 (95% CI 0.82 - 1.05, p<0.001) for SSI-ET, SMD=0.99 (95% CI 0.87-1.11, p<0.001) for B-CBT, and SMD=1.08 (95% CI 0.97 - 1.19, p<0.001) for B-IPT. Finally, standardized symptom reduction for irritability symptoms from baseline to one-month was SMD=0.98 (95% CI 0.85 - 1.10, p<0.001) for SSI-ET, SMD=1.12 (95% CI 1.00-1.25, p<0.001) for B-CBT, and SMD=1.15 (95% CI 1.03 - 1.27, p<0.001) for B-IPT.

**Figure 2.**
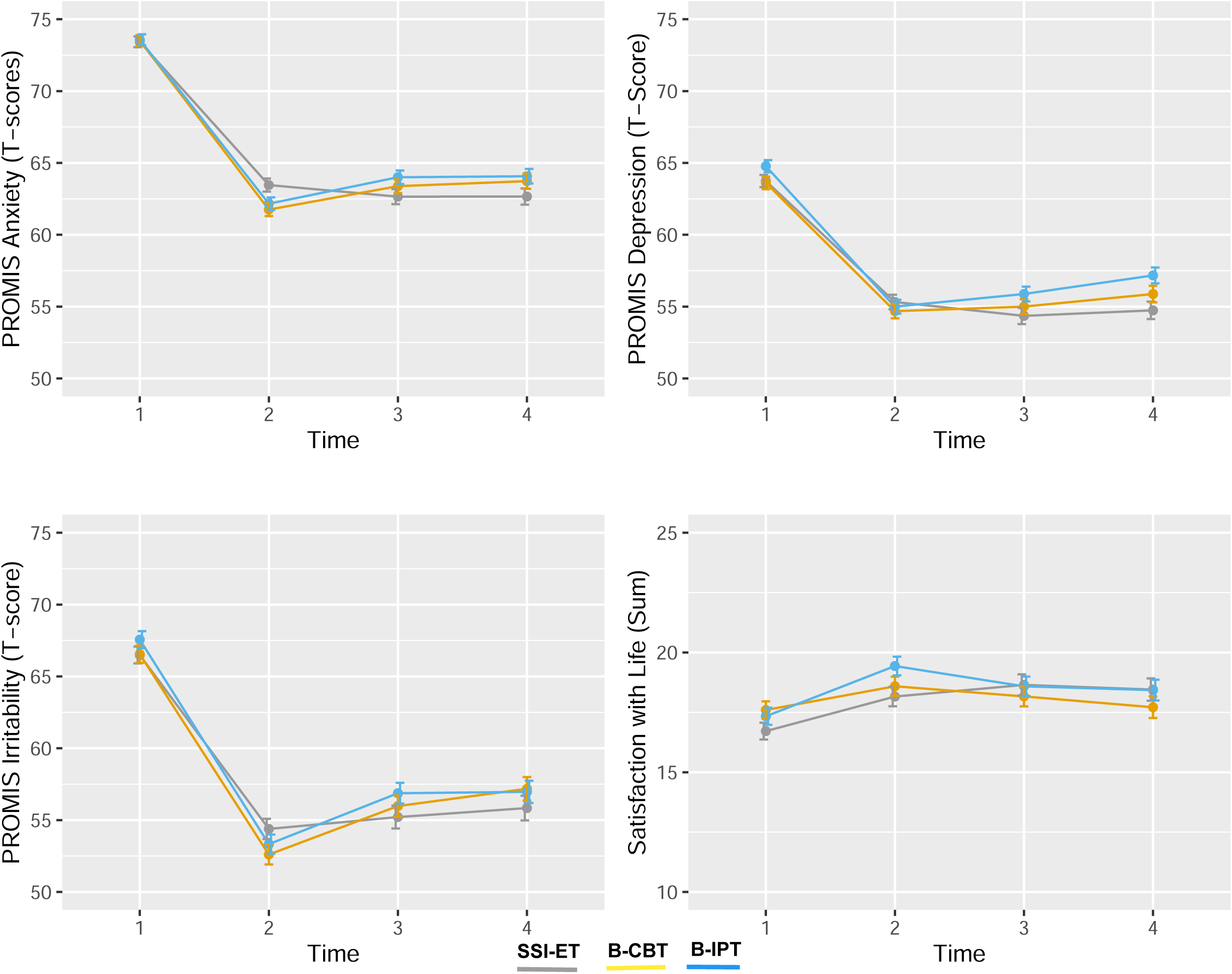
Estimated scores on all secondary outcomes as a function of each timepoint (baseline, one-month, three-months, and six-months follow-up) SSI-ET, single session intervention - enhanced psychoeducation; B-CBT, brief cognitive behavioral telepsychotherapy; B-IPT, brief interpersonal telepsychotherapy

### Time-by-group interactions

We found significant time-by-group interaction for the PROMIS anxiety scores over time (table 2). Pairwise contrasts revealed that both B-CBT and B-IPT were superior to SSI-BT at the one-month assessment, albeit with a small magnitude. The difference between SSI-BT and B-CBT was 1.7 PROMIS T-scores, with an SMD of 0.20 (95% CI 0.05 - 0.34, p=0.007). The difference between SSI-BT and B-IPT was 1.3 PROMIS T-scores, with an SMD of 0.15 (95% CI 0.006 - 0.29), but p-value of 0.04 is not significant if we consider the threshold for pairwise comparisons defined a priori (i.e., p=0.017). None of these differences was maintained at the three- and six-month follow-up assessments. No time-by-group interaction was found in depression and irritability scores; however, a time-by-group interaction was observed for life satisfaction scores. Pairwise contrasts suggested that B-IPT was superior to SSI-BT for life satisfaction scores at one month, with a small non-significant difference of 1.3 points on the life satisfaction scale (SMD 0.19; 95% CI 0.35 - 0.02; p=0.02), again relative to the threshold for pairwise comparisons. Also, these differences were not sustained at the one- or six-month follow-up assessments.

**Table 2.** Linear Mixed Models hypothesis testing, parameter estimates and pairwise comparisons for secondary outcomes.

|  | Model estimated parameters |  |  | Linear mixed effects model | Pairwise comparisons (mean difference, 95% CI) |  |  |
| --- | --- | --- | --- | --- | --- | --- | --- |
| Secondary Outcomes | SSI-ET | B-CBT | B-IPT |  | SSI-ET vs. B-CBT | SSI-ET vs. B-IPT | B-CBT vs. B-IPT |
| **PROMIS Anxiety (EM)** | | | | *Time*<br>$F_{3,2129}=628.3$<br>$p<0.001^*$; | | | |
| Baseline | 73.4 | 73.5 | 73.6 |  | .. | .. | .. |
| One-month (endpoint) | 63.4 | 61.8 | 62.2 | *Group*<br>$F_{2,1106}=0.56$<br>$p=0.573$; | 1.7<br>(0.4 to 3.0)* | 1.3<br>(0.1 to 2.5)* | -0.4<br>(-1.6 to 0.8) |
| Three-months (follow-up) | 62.7 | 63.4 | 64.0 | *Time by Group*<br>$F_{6,2128}=2.94$<br>$p=0.007^*$ | -0.7<br>(-2.1 to 0.7) | -1.3<br>(-2.7 to 0.02) | -0.6<br>(-1.9 to 0.7) |
| Six-months (follow-up) | 62.7 | 63.7 | 64.1 |  | -1.1<br>(-2.6 to 0.5) | -1.4<br>(-2.8 to 0.1) | -0.3<br>(-1.8 to 1.1) |
| **PROMIS Depression (EM)** | | | | *Time*<br>$F_{3,2018}=427.25$<br>$p<0.001^*$; | | | |
| Baseline | 63.7 | 63.6 | 64.8 |  | .. | .. | .. |
| One-month (endpoint) | 55.3 | 54.7 | 55.0 | *Group*<br>$F_{2,1086}=2.93$<br>$p=0.054$; | 0.6<br>(-0.7 to 2.0) | 0.3<br>(-1.0 to 1.7) | -0.3<br>(-1.7 to 1.0) |
| Three-months (follow-up) | 54.4 | 55.0 | 55.9 | *Time by Group*<br>$F_{6,2016}=1.87$,<br>$p=0.081$ | -0.6<br>(-2.1 to 0.8) | -1.5<br>(-3.0 to -0.02) | -0.9<br>(-2.3 to 0.6) |
| Six-months (follow-up) | 54.7 | 55.9 | 57.2 |  | -1.1<br>(-2.8 to 0.5) | -2.4<br>(-4.0 to -0.8) | -1.3<br>(-2.9 to 0.3) |
| **PROMIS Irritability (EM)** | | | | *Time*<br>$F_{3,2032}=378.73$<br>$p<0.001^*$; | | | |
| Baseline | 66.5 | 66.5 | 67.6 |  | .. | .. | .. |
| One-month (endpoint) | 54.4 | 52.6 | 53.3 | *Group*<br>$F_{2,1058}=0.62$<br>$p=0.538$; | 1.78<br>(-0.2 to 3.7) | 1.1<br>(-0.9 to 3.0) | -0.7<br>(-2.6 to 1.2) |
| Three-months (follow-up) | 55.2 | 56.0 | 56.9 | *Time by Group*<br>$F_{6,2030}=1.58$,<br>$p=0.150$ | -0.8<br>(-2.9 to 1.36) | -1.6<br>(-3.8 to 0.4) | -0.9<br>(-2.9 to 1.1) |
| Six-months (follow-up) | 55.8 | 57.2 | 57.0 |  | -1.3<br>(-3.7 to 1.0) | -1.1<br>(-3.4 to 1.1) | 0.2<br>(-2.0 to 2.4) |
| **Satisfaction with life (EM)** | | | | *Time*<br>$F_{3,1903}=20.37$<br>$p<0.001^*$; | | | |
| Baseline | 16.7 | 17.6 | 17.3 |  | .. | .. | .. |
| One-month (endpoint) | 18.2 | 18.6 | 19.4 | *Group*<br>$F_{2,1057}=0.64$<br>$p=0.527$; | -0.4<br>(-1.5 to 0.7) | -1.3<br>(-2.4 to -0.2)* | -0.8<br>(-1.9 to 0.2) |
| Three-months (follow-up) | 18.6 | 18.2 | 18.6 | *Time by Group*<br>$F_{6,1902}=2.46,$<br>$p=0.022^*$ | 0.5<br>(-0.7 to 1.7) | 0.05<br>(-1.1 to 1.2) | -0.4<br>(-1.6 to 0.7) |
| Six-months (follow-up) | 18.5 | 17.7 | 18.4 |  | 0.7<br>(-0.5 to 2.0) | 0.02<br>(-1.2 to 1.3) | -0.7<br>(-1.9 to 0.5) |

PROMIS, Patient-Reported Outcomes Measurement Information System; SSI-ET, single session intervention - enhanced psychoeducation; B-CBT, brief cognitive behavioral telepsychotherapy; B-IPT, brief interpersonal telepsychotherapy; EM, estimated means. \* indicates statistically significant differences.

### Net Promoter Scores

There were significant differences in the probability of participants recommending the treatment among treatment arms. The estimated NPS for each group was 62.4%, 83.7% and 85.6% for SSI-ET, B-CBT, and B-IPT, respectively. Both B-CBT (estimated mean difference=21.3%, p<0.001) and B-IPT (estimated mean difference=23.2%, p<0.001) were superior to SSI-ET in the likelihood of recommending treatment to colleagues. A similar result was also found for the question about satisfaction in each group: 5.48, 5.90, and 6.03 for SSI-ET, B-CBT, and B-IPT, respectively. Both B-CBT (estimated mean difference=0.43, p<0.001) and B-IPT (estimated mean difference= 0.54, p<0.001) were superior to SSI-ET in terms of satisfaction with treatment. An assessment of the open-ended part of the questionnaire revealed that most participants wanted more online sessions.

### Therapists’ fidelity

The supervisors observed nine sessions of SSI-ET, 12 sessions of B-CBT sessions, and eight of B-IPT. The mean fidelity score for therapists in all groups reached almost maximum scores: 4.78 (SD 0.32), 4.89 (SD 0.12), and 4.82 (SD 0.26) for SSI-ET, B-CBT, and B-IPT, respectively, with no significant differences between groups (all p-values>0.05).

### Attrition

The flow diagram in figure 1 illustrates the number of individuals assessed at one-, three-, and six-month follow-up, demonstrating considerable attrition rates. The follow-up rates in the SSI-ET were consistently lower than those in the B-CBT and B-IPT groups across all time points (all p-values<0.05). There was no significant interaction between time and group (p=0.407), indicating that the effect of time on participant loss did not vary between groups.

### Post-hoc equivalence tests

Equivalence tests indicated the observed difference between each pair of interventions were within the equivalence bounds of −15% and 15% for the primary outcome: SSI-ET vs. B-CBT (Z=-3.16, p=0.001), SSI-ET vs. B-IPT (Z=-3.42, p<0.001), and B-CBT vs. B-IPT (Z=3.64, p<0.001).

## DISCUSSION

Our results showed no evidence of superiority between the SSI-TE, B-CBT, and B-IPT interventions. There was a very high symptom reduction in anxiety, depression, and irritability pre-post assessment (SMDs>0.95) in all interventions, that was mostly maintained during the follow-up assessments. There were also considerable estimated response rates after one month (46.5%, 43.7%, and 44.6% for SSI-ET, B-CBT, and B-IPT, respectively), three months (41.5%, 44.6%, and 41.1%), and six months (40.1%, 42.9%, and 33.4%) post-intervention. Thus, contrary to our original hypothesis, we found no evidence to support the use of more complex brief psychotherapies, including components of cognitive-behavioral therapy or interpersonal therapy instead of a single section of enhanced psychoeducation to reduce emotional distress in a context of mental health crisis, such as the one triggered by SARS-CoV-2 pandemic.

Our findings contribute to a growing body of literature on the potential of single session interventions (SSIs) to reduce emotional distress, particularly among healthcare workers facing epidemic crises. SSIs have become a prominent topic in psychological science, with studies demonstrating their high acceptability, utility,^23^ and efficacy for depression^24^ and anxiety,^25^ especially in pediatric populations. There is also robust evidence showing that SSIs are non-inferior to traditional CBT for pain management in adults^26^ and for treating phobias in children.^27^ SSIs are especially attractive for their efficiency, cost-effectiveness, and scalability compared with existing implementation approaches, maintaining positive results during follow-up evaluation.

This study also bolsters the evidence for interventions aligned with Psychological First Aid principles, typically delivered in a single session. Despite widespread dissemination in various crisis settings, the evidence for Psychological First Aid is scant. It is worth noting; however, that while our approach is based on Psychological First Aid principles, it also incorporates elements of psychoeducation, measurement-based care, and risk stratification. The addition of psychoeducational videos enhances the perception of care, providing critical information tailored to patients’ needs during times of crisis and stress.

Another significant aspect of this trial was its transdiagnostic nature. Our study aligns with a contemporary approach of providing therapies that benefit patients with various symptomatic presentations, such as different forms of anxiety and depression,^4,5^ addressing the prevalent observation that comorbidity is common.^28^ Transdiagnostic protocols offer several advantages regarding dissemination and adoption in resource-limited settings.^29^ Our study is among the few that include irritability as a symptom domain, a strength to be emphasized, given the limited therapeutic interventions available for adults with irritability symptoms.^30^ This supports the evidence that irritability is part of the negative affect spectrum, closely linked to anxiety and depression. ^31,32^

Our response rates (i.e. reduction of 50% or more in the primary outcome) are comparable to those reported in other similar trials for depression, for example. In those studies, 41% of those receiving psychotherapy responded to treatment, as compared to 17% for usual care and 16% for waitlist.^33^ Although this comparison is limited by the use of distinct assessment measures, the remission rates were very low for all groups, which contrasts with the literature that shows that around a third of patients remit after therapy for depression.^33^ This indicates that, although the interventions likely led to a substantial symptom reduction, many participants continued to experience symptoms at subthreshold levels. Thus, pairing this intervention with long-term treatments for participants who continue to present symptoms after brief interventions and remain motivated for psychotherapy might be a required strategy for improving remission rates. This is also consistent with the results showing higher satisfaction rates in participants receiving four sessions of therapy. Besides this, it is possible that SSI increased openness or motivation to engage with additional mental health services. A recent randomized trial showed that one single session embedded within social media platforms dramatically increased young people’s odds of accessing mental health resources in moments of crisis.^34^ Unfortunately, we did not measure additional mental health engagement in our study and future clinical trials could significantly contribute to this discussion by including assessments of motivation in participants.

Regarding treatment adherence, we could identify that both B-CBT and B-IPT had similar rates, whereas the SSI-ET had a 100% of adherence due to the duration of the intervention. However, the follow-up response shows that those who received four sessions were more prone to achieve response during the follow-up, leading to a high attrition rate in our analysis. Our treatment adherence and dropout during the follow-up period is consistent with what has been previously reported in literature, even though our study was online and covered a continental country.

This study has some limitations that should be acknowledged. First, there was a high attrition rate, particularly at the three- and six-month follow-ups. Although this is consistent with large pragmatic trials, our estimates might be biased due to the lack of representativeness of the original sample. We minimized these biases using appropriate statistical methods that consider all available data to provide adjusted estimates. Second, this trial was conducted entirely in a virtual environment, relying heavily on self-reports and clinical assessments, without formal diagnostic interviews or clinician-performed assessments with standard instruments. While this reflects real-world conditions, especially in lower-middle income countries or countries with large geographical areas, such as Brazil, it limits comparability with other clinical studies. Third, the absence of a non-active comparison condition restricts our ability to infer the effectiveness of each protocol. Although scientifically valuable, the high level of emotional distress among healthcare professionals in a country with numerous COVID-19 cases precluded the use of a non-active intervention from an ethical perspective. Besides this, PROMIS scales were adapted for the Brazilian Portuguese, but have not been normatized for this population.

Thus, the T-scores used in this study derived from a different population. Another important aspect to consider is that since the trial occurred from May 2020 to December 2021, this period includes distinct waves, making our sample particularly heterogeneous. Anecdotally, therapists described that the first cases were represented by more acute situations; whereas last waves were more represented by burnout and exacerbation of chronic conditions. Finally, the specialized interventions were still brief, with only four sessions, and more extensive interventions could have resulted in a significant difference when compared with the SSI-ET.

This study is one of the largest of its kind to support a very simple SSI-ET intervention. Accordingly, future studies exploring the effectiveness of brief, transdiagnostic, and less specialized interventions may improve the evidence in favor of more cost-effective and scalable interventions, aspects that are crucial for lower-middle income countries. Predicated on the essential types of support that should be provided in a crisis, it involves actively listening to individuals experiencing emotional distress, validating their feelings, connecting with their experiences, providing empathic responses for each identified problem, assessing their symptoms and coping mechanisms, offering education on helpful strategies, and tracking progress with empirically validated assessments. Our study suggests that these simple elements may be sufficient during crises and do not support the additional benefits of adding brief CBT- and IPT-specific strategies for adults experiencing significant emotional distress in a crisis.

## Contributors

GAS and CBD conceived and designed the study. GAS and CBD obtained funding for the trial. GAS, MAC, LS, AS, NPG, and LHS provided administrative, technical, or material support. GAS developed the statistical analysis plan. GAS, AS, and NPG conducted the statistical analysis. GAS, PC, DSP, ARB, CHK, MPAF, GGM, and CBD supervised the conduction of the trial and production of this Article. All authors drafted and critically revised the content of this Article. All authors have read and approved the final manuscript, confirm that they had full access to the data, and accept responsibility to submit for publication.

## Declaration of interests

All authors have completed and submitted the ICMJE Form for Disclosure of Potential Conflicts of Interest. All authors declare there is no conflict of interest concerning the scientific content and the ideas expressed in this paper.

## Data sharing

Individual participant data that underlie the results reported in this article, after de-identification (text, tables, figures, and appendices) and the study protocol will be available immediately following publication with no end date. Researchers who provide a methodologically sound proposal may gain access to the database to achieve aims in the approved proposal. Proposals should be directed to and, to gain access, data requestors will need to sign a data access agreement. After 36 months, data will be available for 5 years at our university’s data warehouse (https://lume.ufrgs.br), but without investigator support other than deposited metadata.

## Acknowledgment

This study was financed in part by the Ministry of Health of Brazil through, the decentralized execution term No. 16/2020, Coordenação de Aperfeiçoamento de Pessoal de Nível Superior (CAPES), Finance Code 001, Conselho Nacional de Desenvolvimento Científico e Tecnológico (CNPq - 001), Brazilian federal government agencies, and Fundo de Incentivo à Pesquisa/Hospital de Clínicas de Porto Alegre (FIPE/HCPA - 001) – Brazil. The funders of the study had no role in the design or conduct of the study; collection, management, analysis or interpretation of the data; preparation, review or approval of the manuscript; or decision to submit the manuscript for publication. The views expressed are those of the authors and not necessarily those of the Ministry of Health of Brazil, CAPES, CNPq, and FIPE/HCPA. This project was only possible due to the effort and collaboration of many people and institutions; therefore, we would like to thank our collaborators from Hospital de Clínicas de Porto Alegre, Pontifícia Universidade Católica do Rio Grande do Sul, Universidade Federal de Ciências da Saúde de Porto Alegre, Universidade de São Paulo, Universidade Federal de São Paulo, Pan American Health Organization/WHO, Sociedade Brasileira de Psicologia, Associação de Psiquiatria do Rio Grande do Sul, Universidade Federal do Paraná, Universidade Estadual de Campinas, Centro de Estudos em Psiquiatria Integrada, Liga de Psiquiatria e Saúde Mental da UFCSPA/UFRGS, The Funn Club, Yoho Arsenal, and Agência LVL Funding.

## Notes

### Competing Interest Statement

The authors have declared no competing interest.

### Clinical Trial

ClinicalTrials.gov, NCT04635618

### Funding Statement

This study was funded in part by the Ministry of Health of Brazil through, the decentralized execution term No. 16/2020, Coordenacao de Aperfeicoamento de Pessoal de Nivel Superior (CAPES), Finance Code 001, Conselho Nacional de Desenvolvimento Cientifico e Tecnologico (CNPq - 001), Brazilian federal government agencies, and Fundo de Incentivo a Pesquisa/Hospital de Clinicas de Porto Alegre (FIPE/HCPA - 001), Brazil.

### Author Declarations

The trial protocol was approved by the National Research Ethics Commission (CONEP).

