## appendix for "Brief remote psychological treatments for healthcare workers with emotional distress during the SARS-CoV-2 pandemic: a randomized clinical trial"

**Supplementary Appendix**

**Contents**

**Appendix 1: CONSORT checklist (pg. 2)**

**Appendix 2: Protocol of the single session intervention-enhanced telepsychoeducation group (SSI-ET) (pg. 4)**

**Appendix 3: Protocol of the brief cognitive-behavioral telepsychotherapy group (B-CBT) (pg. 6)**

**Appendix 4: Protocol of the brief interpersonal telepsychotherapy group (B-IPT) (pg. 8)**

**Appendix 5: Sociodemographic description of groups (pg. 10)**

**Appendix 1: CONSORT checklist**

| **Section/Topic** | **Item No** | **Checklist item** | **Reported on page No** |
| --- | --- | --- | --- |
| **Title and abstract** | | | |
|  | 1a | Identification as a randomised trial in the title | 1 |
|  | 1b | Structured summary of trial design, methods, results, and conclusions (for specific guidance see CONSORT for abstracts) | 3 |
| **Introduction** | | | |
| Background and objectives | 2a | Scientific background and explanation of rationale | 4 |
|  | 2b | Specific objectives or hypotheses | 4 |
| **Methods** | | | |
| Trial design | 3a | Description of trial design (such as parallel, factorial) including allocation ratio | 5 |
|  | 3b | Important changes to methods after trial commencement (such as eligibility criteria), with reasons | 5 |
| Participants | 4a | Eligibility criteria for participants | 5 |
|  | 4b | Settings and locations where the data were collected | 5 |
| Interventions | 5 | The interventions for each group with sufficient details to allow replication, including how and when they were actually administered | 6-8; appendix |
| Outcomes | 6a | Completely defined pre-specified primary and secondary outcome measures, including how and when they were assessed | 8-9 |
|  | 6b | Any changes to trial outcomes after the trial commenced, with reasons | 8-9 |
| Sample size | 7a | How sample size was determined | 9 |
|  | 7b | When applicable, explanation of any interim analyses and stopping guidelines | 10 |
| Randomisation: |  |  | 6 |
| Sequence generation | 8a | Method used to generate the random allocation sequence |  |
|  | 8b | Type of randomisation; details of any restriction (such as blocking and block size) | 6 |
| Allocation concealment mechanism | 9 | Mechanism used to implement the random allocation sequence (such as sequentially numbered containers), describing any steps taken to conceal the sequence until interventions were assigned | 6 |
| Implementation | 10 | Who generated the random allocation sequence, who enrolled participants, and who assigned participants to interventions | 6 |
| Blinding | 11a | If done, who was blinded after assignment to interventions (for example, participants, care providers, those assessing outcomes) and how | 5 |
|  | 11b | If relevant, description of the similarity of interventions | 6-8 |
| Statistical methods | 12a | Statistical methods used to compare groups for primary and secondary outcomes | 10 |
|  | 12b | Methods for additional analyses, such as subgroup analyses and adjusted analyses | 10 |
| **Results** | | | |
| Participant flow (a diagram is strongly recommended) | 13a | For each group, the numbers of participants who were randomly assigned, received intended treatment, and were analysed for the primary outcome | 11; Fig. 1 |
|  | 13b | For each group, losses and exclusions after randomisation, together with reasons | Fig. 1 |
| Recruitment | 14a | Dates defining the periods of recruitment and follow-up | 11 |
|  | 14b | Why the trial ended or was stopped | 10 |
| Baseline data | 15 | A table showing baseline demographic and clinical characteristics for each group | Table 1 |
| Numbers analysed | 16 | For each group, number of participants (denominator) included in each analysis and whether the analysis was by original assigned groups | Table 1 |
| Outcomes and estimation | 17a | For each primary and secondary outcome, results for each group, and the estimated effect size and its precision (such as 95% confidence interval) | 11-13; Table 2 |
|  | 17b | For binary outcomes, presentation of both absolute and relative effect sizes is recommended | 11-13; Table 2 |
| Ancillary analyses | 18 | Results of any other analyses performed, including subgroup analyses and adjusted analyses, distinguishing pre-specified from exploratory | 11-13; Table 2 |
| Harms | 19 | All important harms or unintended effects in each group (for specific guidance see CONSORT for harms) | 13 |
| **Discussion** | | | |
| Limitations | 20 | Trial limitations, addressing sources of potential bias, imprecision, and, if relevant, multiplicity of analyses | 15 |
| Generalisability | 21 | Generalisability (external validity, applicability) of the trial findings | 14-15 |
| Interpretation | 22 | Interpretation consistent with results, balancing benefits and harms, and considering other relevant evidence | 14-15 |
| **Other information** | | |  |
| Registration | 23 | Registration number and name of trial registry | 10 |
| Protocol | 24 | Where the full trial protocol can be accessed, if available | 16 |
| Funding | 25 | Sources of funding and other support (such as supply of drugs), role of funders | 16 |

**Appendix 2: Protocol of the single session intervention-enhanced telepsychoeducation group (SSI-ET)**

The enhanced psychoeducation protocol is a one-session intervention based on supportive principles delivered by video call. The general principle is assisting participants in stressful situations using components of psychological first aid. Therapists are oriented to respect participants' safety, dignity, and rights, adapt the intervention to cultural issues, and encourage participants to develop autonomy and self-efficacy. Moreover, the therapists should listen carefully and empathetically to the participants and create a welcoming, gentle, and non-judgmental environment where participants feel comfortable and confident to express their thoughts, concerns, and emotions. Therapists should help participants resolve problems without harming them, strengthen their support network, and develop the ability to feel safe, close to people, calm, and hopeful, as well as ensure social, physical, and emotional support. Improving the coping of the participants, generating greater self-confidence, improving satisfaction, and reducing anxiety, depression, and emotional pain are the main targets of the intervention. The principles of observing, listening, and connecting guide the entire session.

The psychoeducation session has four objectives: (1) offering a safe environment for the participant to express their emotions, and thoughts and share their experiences (10 to 30 minutes); (2) reviewing with the participant the symptomatic assessment which was self-rated by the participant in the project' registration, helping him/her on the areas of greatest concern (5 to 15 minutes); (3) planning lifestyle changes, promoting protective factors and reducing risk factors for mental health; (4) planning the use of psychoeducation videos that will be used over the next four weeks (in the intervention with support videos; ten minutes).

Empathetic listening is the essential component of the entire session, and the main technique adopted in this modality. Listening carefully to the participant's demand, validating emotions and efforts, demonstrating the therapist is attentive, and using verbal and non-verbal components of empathy are the basis of empathetic listening. Other interviewing techniques are encouraged: a. facilitation (refers to the posture, actions, or words that encourage the participant to communicate); b. echo (repetition of participant's words encouraging him/her to express feelings); c. empathic responses; d. validation (the therapist should look for what can be validated in the participant's speech); e. tranquilization; f. summarizing the participant's narrative; g. highlighting transitions in the sessions. h. adapting the language for the participant.

In the intervention with supportive videos, the videos are personalized based on the demands identified in the questionnaire answered by the participant and during the session. The therapists selected eight videos, and sent them to the participant for four weeks (two videos per week), considering the best order and moment to send them (during day-time). Moreover, the therapist was available during this period to answer participants’ messages by phone chat. The following themes comprise each video: 1. SARS-CoV2: How to protect yourself from SARS-CoV2 when you arrive at your home?; 2. Fear of contagion; 3. Normal Anxiety vs. Excessive Anxiety; 4. Sadness vs. Depression; 5. Anger vs. irritability; 6. Burnout: What is Burnout?; 7. Stress and acute reaction to stress; 8. Sleep: Sleep hygiene; 9. Food: Healthy eating and mental health; 10. Exercise: Exercise and mental health; 11. Substances; 12. News: How to protect yourself from excessive news exposure? 13. Social Networks: Excessive use of social networks; 14. Children: How to deal with your children in quarantine times?; 15. Elderly: How to deal with elderly people in quarantine times? 16. Social support: How to stay connected in quarantine times?

**Appendix 3: Protocol of the brief cognitive-behavioral telepsychotherapy group (B-CBT)**

The cognitive-behavioral psychotherapy protocol is based on cognitive-cognitive-behavioral theory and delivered by video call. This brief intervention consists of 4 weeks and focuses on the most frequent clinical symptoms and common mental disorders. This strategy also aims to provide resources to health professionals to cope with stress. The techniques included were based on previous studies that showed effectiveness in brief interventions. This protocol used a transdiagnostic approach based on the Unified Protocol for Transdiagnostic Treatment, an intervention designed to address neuroticism, the core temperament associated with anxious and depressive disorders. Due to the pandemic context, we also included techniques to deal with insomnia, acute stress, emotional distress, and irritability. Each session was followed with two booster videos sent by text messages.

The protocol is carried out in four remote sessions with the same therapist using a video call with Google Meet. The protocol comprises an initial assessment session and three cognitive-behavioral therapy sessions. In summary, session one was composed of a clinical evaluation and contained components of psychological first aid and the use of personalized remote psychoeducational videos. The objectives of this session were the following: (1) to offer a safe place for the participants to express and share their emotions; (2) to assist the participant in urgent demands; (3) to give feedback on the symptomatic evaluation assessment; (4) to guide the participant on the protocol sessions, establishing the therapy time and duration, and (5) to plan the two videos with personalized psychoeducational resources. Session two comprises psychoeducation and techniques for the management of irritability and anxiety. The components of this session are the following: psychoeducation on the cognitive model, acceptance of emotions in a non-judgmental manner, diaphragmatic breathing and mindfulness practice. Session three deals predominantly with cognitive restructuring components. In this session, the therapist approaches perceptions of self-competence, suffering, excessive guilt and stigma. The cognitive model is reviewed looking for flexible thinking, alternative thoughts and reassignment techniques. And finally, session four focuses on behavioral strategies and avoidance. This session emphasizes emotional self-regulation. The problem-solving technique is contextualized within the participant's demand, and strategies are focused on the solution or acceptance. The therapist detects dysfunctional areas and encourages behavioral activation as a way of promoting mental health. Connection with the network, friends, or family, organizing rest and pleasure activities are reinforced. Finally, the importance of developing strategies to prevent relapse are addressed.

During the intersession period, the therapists were responsible for sending two videos per week, totaling eight videos over the four-week treatment duration. These videos were carefully selected in a personalized manner to meet the individual's needs. The selection process involved incorporating videos available within the psychoeducation group, as well as specific videos tailored to CBT.

**Appendix 4: Protocol of the brief interpersonal telepsychotherapy group (B-IPT)**

Interpersonal psychotherapy (IPT) is an adaptation of interpersonal therapy to be applied in four sessions in the context of the COVID-19 Pandemic. Tele B-IPT model was based on The Edinburgh Early Intervention Model: Psychological First Aid & Abbreviated Interpersonal Psychotherapy Adapted for COVID-19 and interpersonal counseling (IPC) for depression in primary care. The B-IPT was initially developed for health professionals involved in caring for patients with COVID-19 and then extended to other professionals directly involved with the pandemic (e.g. teachers and security professionals).

IPT is a form of brief psychotherapy, originally developed for depression, which seeks to understand the interpersonal and social context in which the symptoms arise and how they are related. IPT assumes that interpersonal problems and depression are closely associated and influence each other (interpersonal problems can lead to depression and depression can lead to interpersonal problems). Progressively, IPT has also been studied in other disorders, such as anxiety disorders, trauma, and emotional dysregulation. It is performed by the same therapist, in four sessions carried out by a phone call or preferentially through video call. Remote sessions include an initial assessment and psychoeducation session (session one) and three sessions (sessions two to four) with interpersonal therapy techniques. Remote sessions are preferably scheduled over four weeks. IPT-B, with its problem areas related to grief, interpersonal disputes, and role transitions, proved to be an appropriate intervention model for the crisis context, particularly for health professionals who dealt with the health demands of the pandemic.

The objective of session one is to establish a therapeutic alliance, through empathetic listening with elements of first psychological care. In addition, risk assessment, severity and syndromic diagnosis are carried out. In session one, we also look for the psychosocial and interpersonal context in which the participant is living, to understand their relationship with the symptoms presented. Then, the interpersonal inventory is elaborated, which is a mapping of the participant's relationships (past and current), understanding who are the people with whom he/she relates, what is the level of intimacy that the participant has with them, what are the mutual expectations and how much does he/she count on these people as a support network. In the context of the pandemic, it is essential to understand whether the current support network has been modified and the degree of this change, in addition to establishing whether this change is related to the participant's symptoms. Also, within the first session, it is sought to determine which problem area will be chosen as the focus of the other sessions: grief, transition of roles, or interpersonal dispute.

The grieving problem area refers to the loss of an important person in the participant's life. The role transition problem area refers to some change that occurred in the participant's life and the need to adapt to this change. In the context of COVID-19, this is a frequent problem area and may involve difficulties in dealing with the new situation at work, experiencing social isolation, and having to take all precautions related to non-contamination, among others. The pandemic requires a transition of roles since it requires profound changes in various areas of life such as social isolation, changes in routine, or care to avoid contamination of professionals. It is necessary to identify, at this stage, whether and which of these role transitions are related to the symptoms of the participant in question. The interpersonal dispute problem area is chosen when a participant's conflict with a family member, co-worker or significant person is occurring.

In sessions two and three the initial symptoms of emotional distress are reviewed, relating them to the interpersonal context. The problem area identified in session one will be worked on in the following sessions, maintaining continuity between sessions and deepening strategies to deal with the situation in focus. The therapist makes use of the entire repertoire of IPT techniques, such as non-directive exploration, directive elicitation, clarifications, communication analysis, decision analysis, staging, counseling, and mentalization.

In session four, in addition to the usual symptom review, the end of treatment is explicitly discussed. The therapist helps the participant to recognize his/her competence through the review of the interpersonal network, reviews the course of treatment and the progress of the participant and assesses early warning signs, and discusses future alternatives.

During the intersession period, the therapists were responsible for sending two videos per week, totaling eight videos over the four-week treatment duration. These videos were carefully selected in a personalized manner to cater to the individual's needs. The selection process involved incorporating videos available within the psychoeducation group, as well as specific videos tailored to IPT.

| **Appendix 5: Sociodemographic description of groups** | | | |
| --- | --- | --- | --- |
|  | **SSI-ET (**n=342) | **B-CBT** (n=323) | **B-IPT (**n=334) |
| **Country region** |  |  |  |
| South | 114 (33.3%) | 102 (31.6%) | 110 (32.9%) |
| Southeast | 131 (38.3%) | 122 (37.8%) | 142 (42.5%) |
| Midwest | 32 (9.4%) | 25 (7.7%) | 27 (8.1%) |
| Northeast | 56 (16.4%) | 66 (20.4%) | 44 (13.2%) |
| North | 9 (2.6%) | 9 (2.5%) | 11 (3.3%) |
| **Profession** |  |  |  |
| Physicians | 16 (4.7%) | 15 (4.6%) | 20 (6.0%) |
| Nurses | 145 (42.4%) | 146 (45.2%) | 147 (44.0%) |
| Nursing technicians | 12 (3.5%) | 17 (5.3%) | 9 (2.7%) |
| Interns | 5 (1.5%) | 5 (1.5%) | 9 (2.7%) |
| Administrative professionals | 37 (10.8%) | 38 (11.8%) | 36 (10.8%) |
| Community agents | 36 (10.5%) | 31 (9.6%) | 34 (10.2%) |
| Other professionals | 91 (26.6%) | 71 (5.3%) | 79 (23.7%) |
| **Any treatment for mental disorders (yes)** | 114 (33.3%) | 116 (36.1%) | 121 (36.3%) |
| **Pharmacological treatment (yes)** | 160 (46.8%) | 157 (48.9%) | 152 (45.6%) |
| **Psychological treatment (yes)** | 34 (9.9%) | 42 (13.1%) | 37 (11.1%) |
| SSI-ET, single session intervention - enhanced psychoeducation; B-CBT, brief cognitive behavioral telepsychotherapy; B-IPT, brief interpersonal telepsychotherapy. | | | |
